# Descriptive epidemiology of physical activity energy expenditure in UK adults. The Fenland Study

**DOI:** 10.1101/19003442

**Authors:** Tim Lindsay, Kate Westgate, Katrien Wijndaele, Stefanie Hollidge, Nicola Kerrison, Nita Forouhi, Simon Griffin, Nick Wareham, Søren Brage

## Abstract

**Background:** Physical activity (PA) plays a role in the prevention of a range of diseases including obesity and cardiometabolic disorders. Large population-based descriptive studies of PA, incorporating precise measurement, are needed to understand the relative burden of insufficient PA levels and to inform the tailoring of interventions. Combined heart and movement sensing enables the study of physical activity energy expenditure (PAEE) and intensity distribution. We aimed to describe the sociodemographic correlates of PAEE and moderate-to-vigorous physical activity (MVPA) in UK adults.

**Methods:** The Fenland study is a population-based cohort study of 12,435 adults aged 29–64 years-old in Cambridgeshire, UK. Following individual calibration (treadmill), participants wore a combined heart rate and movement sensor continuously for 6 days in free-living, from which we derived PAEE (kJ•day^-1^•kg^-1^) and time in MVPA (>3 & >4 METs) in bouts greater than 1 minute and 10 minutes. Socio-demographic information was self-reported. Stratum-specific summary statistics and multivariable analyses were performed.

**Results:** Women accumulated a mean(sd) 50(20) kJ•day^-1^•kg^-1^ of PAEE, and 83(67) and 33(39) minutes•day^-1^ of 1-min bouted and 10-min bouted MVPA respectively. By contrast, men recorded 59(23) kJ•day^-1^•kg^-1^, 124(84) and 60(58) minutes•day^-1^. Age and BMI were also important correlates of PA. Association with age was inverse in both sexes, more strongly so for PAEE than MVPA. Obese individuals accumulated less PA than their normal-weight counterparts whether considering PAEE or allometrically-scaled PAEE (-10 kJ•day^-1^•kg^-1^ vs -15 kJ•day^-1^•kg^-2/3^ in men). Higher income and manual work were associated with higher PA; manual workers recorded 13-16 kJ•kg^-1^•day^-1^ more PAEE than sedentary counterparts. Overall, 86% of women and 96% of men accumulated 21.4 min•day^-1^ of MVPA (>3 METs) on average (150 minutes per week). These values were 49% and 74% if only considering bouts >10 min (15% and 31% for >4 METs).

**Conclusions:** PA varied by age, sex and BMI, and was higher in manual workers and those with higher incomes. Light physical activity was the main driver of PAEE; a component of PA that is currently not quantified as a target in UK guidelines.

## Background

Physical activity (PA) plays an important role in the prevention of a range of diseases including obesity and cardiometabolic disorders [1–3]. It is hence an important behavioural target for public health interventions, and guidelines describing desired levels of PA have been proposed [4, 5]. In order to assess the population burden of insufficient levels of PA, and develop effective, tailored interventions, it is important to describe physical activity levels and examine the socio-demographic correlates.

Human behaviour occurs across an intensity spectrum ranging from sleep and sedentary behaviour (SS), to light physical activity (LPA), moderate physical activity (MPA) and vigorous physical activity (VPA). Typically, these intensities are grouped according to a metabolic equivalent of task (METs), e.g. LPA considered as 1.5-3 METs, MPA as 3-6 METs, and VPA as greater than 6 METs, although such classification is far from universal [6]. Recent updates in physical activity guidelines emphasize the importance of moderate-to-vigorous PA (MVPA) but increasingly recognise the importance of all subcomponents of the entire intensity spectrum [4]. Moreover, the 2018 guidelines for Americans no longer include the requirement for activity to occur in bouts of at least 10-min duration. Whilst other countries may adopt this definition, it is far from universal and the consequences of such reclassification in terms of activity levels in different socio-demographic groups are not well documented with objective measurements in large cohorts.

Objectively measured PAEE of smaller British cohorts has been reported, including a nationally representative sample of children and adults measured with the doubly labelled water method [7]; however, this method can only assess total volume of PAEE, and the sample was too small (n = 770) to describe socio-demographic differences. Two other British cohort studies have described PAEE and its underlying intensity distribution, one in adolescents aged 16y and one in older adults aged between 60 and 64y, respectively [8, 9]. The descriptive epidemiology of PAEE and intensity in younger to middle-aged UK adults has not yet been reported. To fill this knowledge gap, we used data from the Fenland cohort, an ongoing population-based observational study of 12,435 adults aged 29-64 years of age, residing in Cambridgeshire, UK. We aimed to describe the objectively measured levels of PAEE and underlying intensity patterns by socio-demographic characteristics.

## Methods

### Study population

Participants born between 1950 and 1975 were recruited to the Fenland Study from general practice lists between 2005 and 2015. Exclusion criteria included pregnancy, physician-diagnosed diabetes, inability to walk unaided, psychosis, and terminal illness. In total, 12,435 participants, aged 29–64 years old, were enrolled and attended one of three clinical research facilities (Ely, Cambridge, and Wisbech) after an overnight fast. All participants provided written informed consent and the study was approved by the local ethics committee (NRES Committee – East of England Cambridge Central) and performed in accordance with the Declaration of Helsinki. In addition, approval was granted to compare general practice-held information (age, sex, height, weight, smoking, alcohol consumption, area deprivation score) for participants with the overall eligible sample (under UK Section 251 legislation).

### Anthropometry and other clinical measures

Height was measured with a rigid stadiometer (SECA 240; Seca, Birmingham, UK) and weight was measured in light clothing with calibrated scales (TANITA model BC-418 MA; Tanita, Tokyo, Japan). Seated blood pressure was measured with an automated sphygmomanometer (Omron, 705CP-II) and a 12-lead ECG (Seca CT6i) was obtained during supine rest. Participants were cleared for submaximal exercise testing if blood pressure was <180/110 mm Hg, no serious anomalies were observed on the ECG, alongside appropriate responses to medical screening questions and a review of medication (e.g. high-dose betablocker use were excluded from the treadmill test).

### Objective physical activity assessment

Participants were fitted with a combined heart rate and uniaxial movement sensor (Actiheart, CamNtech, Papworth, UK), attached to the chest with standard ECG electrodes [10]. Participants had their heart rate measured continuously during a 6-min supine rest test and then underwent a submaximal treadmill test consisting of 9-min of walking on the flat with increasing speed, 6-min walking at increased gradient, and 5-min of jogging on the flat as previously described [11]. Heart rate was measured continuously and the test was terminated when heart rate reached 90% of age-predicted maximum [12], or had been above 80% for longer than 2-min, or the participant requested to stop. At the end of the clinical visit, participants were asked to wear the heart rate and movement sensor, initialised to collect date at 1-min resolution, for the following 6 days [10], and to return the monitor by freepost. Participants were advised that the device was waterproof and should be worn continuously, including during showering, water-based activities, and sleeping, whilst continuing with their usual activities. It could be removed to change electrodes, spare sets of which were provided.

Following pre-processing of the free-living heart rate data to eliminate potential noise [13], heart rate was individually calibrated using parameters obtained from the treadmill test [11] and combined with acceleration in a branched equation model [14] to calculate instantaneous PAEE (J•kg^-1^•min^-1^); this measure of intensity agrees well with intensity measured using indirect calorimetry [15, 16]. For participants without sufficient treadmill data for individual calibration (n = 468 of 12002), an age-, sex-, and sleeping heart rate (SHR) adjusted group calibration was used, derived on all other participants as follows:

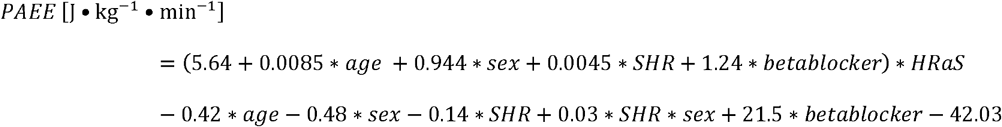

(age in years, sex coded as 1 for men and 0 for women, SHR in beats per minute, heart rate above sleep (HRaS) in beats per minute, and betablocker coded as 0 or 1 if the participant was taking betablocker medication).

Intensity was expressed in standard metabolic equivalents (METs), using 1 MET = 71 J•min^-1^•kg^-1^ (∼3.5 ml O_2_•min^-1^•kg^-1^).

Segments of data with continuous zero acceleration lasting ≥90 minutes were classified as ‘non-wear’ if also accompanied by non-physiological HR data, i.e. consistently high Bayesian error [17]. Average daily PAEE (in kJ•day^-1^•kg^-1^) and time spent at multiple intensity levels (up to 10 METs) and in at least moderate intensity bouts was summarised, whilst minimising potential diurnal bias by imbalance of wear time [18]. This method has been successfully validated against PAEE from doubly-labelled water in UK men and women [19]. Time and energy spent at specific intensity levels were grouped together and defined as sedentary/sleep (SS), light physical activity (LPA), moderate physical activity (MPA) and vigorous physical activity (MPA). Activity bouts of at least 3 and 4 MET (moderate) intensity were calculated in durations ranging from 1 to 10 mins. For the primary analysis SS was defined as < 1.5 METs, LPA as 1.5-3 METs, MPA as 3-6 METs, and VPA as >6 MET. Sensitivity analyses were conducted using LPA as 1.5-4 METs, MPA as 4-7 METs, and VPA as >7 METs, and allometrically scaled PAEE (in kJ•day^-1^•kg^-2/3^) as previously described [20, 21].

For the present analysis, participants were excluded (n=351) if they had worn their sensors for <72 hours overall, or <8 hours accumulative wear for each quadrant of the day (3am to 9am, 9am to 3pm, 3pm to 9pm, 9pm to 3am). Furthermore, activity records were excluded if they did not measure 0 m•s^-2^ at some point (indicating no movement) during the monitoring period to safeguard the analysis from technical accelerometer errors (n=82).

### Covariates

Socio-demographic and lifestyle behavioural information was collected using self-report. This included age, sex, ethnicity, work-type/status (sedentary, standing, manual, retired, unemployed), marital status (single, married/cohabiting, widowed/separated/divorced), education (compulsory, further – A-level/apprenticeship/sub-degree level, higher – degree level or above), household income level (<£20,000, £20,000 - £40,000, >£40,000), smoking status (never, former, current) and alcohol intake (units/week). In addition, location (Cambridge, Ely, Wisbech) and season of physical activity measurement was considered (coded as two orthogonal sine functions; “Winter” peaking at 1 on January 1^st^ and reaching a minimum of -1 on July 1^st^, and Spring peaking at 1 on April 1^st^ and reaching a minimum of -1 on October 1^st^).

### Statistics

All analyses were sex-stratified. We report medians (inter-quartile ranges) or means (standard deviations) for descriptive purposes for continuous variables and proportions for categorical variables. We performed linear test for trend of activity differences for ordinal covariates and likelihood-ratio tests for categorical covariates. We used sex-stratified multivariable linear regression to model the independent associations of activity outcomes with age, BMI, education level, work-type, income, marital status, test site, smoking status, ethnicity, and season of activity measurement. For this analysis, missing data in categorical variables were coded as a separate category.

Using all available information from the general practices, we compared all invited vs all participating by two-sample, unpaired t-test for continuous variables and chi square test for categorical variables. All statistical analyses were performed using Stata/SE version 14.

## Results

A total of 46,024 individuals were invited, of whom 12,435 (27% response rate) agreed to participate. General practice information was obtained on 45,043 individuals including 12,145 study participants; one practice could not provide any standard information (n=180), and a few additional participants did not consent to this linkage, had no valid NHS number or their GP surgery information was missing (n=110). Compared to the overall sampling frame participants were approximately 18 months older, had lower BMI [0.2 (men); 0.8 (women) kg/m^2^ lower], were less likely to smoke, and had marginally lower deprivation scores. Although there was a higher prevalence of alcohol drinkers, participants drank fewer units per week (Supplementary Table 1).

Of the 12,435 participants, 12,002 (91% White) met the combined sensing inclusion criteria for the present analysis, more women (n=6,428) than men (n=5,574). This subsample did not significantly differ from the full Fenland sample, with the exception of men being 0.1 years younger (p=0.02).

Mean (SD) PAEE for women was 50 (20) kJ•day^-1^•kg^-1^ compared to 59 (23) kJ•day^-1^•kg^-1^ for men (Table 1). Women recorded an average of 83 (67) minutes per day of MVPA, of which 33 (39) min/day occurred in bouts of 10-min or longer, whereas men recorded 124 (85) and 60 (58) min/day, respectively. Of note, despite the differences in PAEE and MVPA as measured by combined sensing, women and men did not differ by chest acceleration. Further participant characteristics are listed in Table 1 and univariable analysis by category in Supplementary Table 2.

**Table 1:** Participant characteristics. The Fenland Study 2005 to 2015.

|  |  | Women<br>n = 6428 |  | Men<br>n = 5574 |  |
| --- | --- | --- | --- | --- | --- |
| **Age (years)** |  | 48.7 | 7.4 | 48.6 | 7.6 |
| **BMI (kg/m<sup>2</sup>)** |  | 26.5 | 5.3 | 27.3 | 4.1 |
| **Ethnicity** |  |  |  |  |  |
|  | White | 5954 | 92.6 | 5165 | 92.7 |
|  | South Asian | 74 | 1.2 | 75 | 1.4 |
|  | Black | 30 | 0.5 | 31 | 0.6 |
|  | East Asian | 44 | 0.7 | 20 | 0.4 |
|  | Others or unknown | 326 | 5.0 | 283 | 5.1 |
| **Education level** |  |  |  |  |  |
|  | Basic | 1442 | 22.4 | 1005 | 18.0 |
|  | Further | 2923 | 45.5 | 2601 | 46.7 |
|  | Higher | 2063 | 32.1 | 1968 | 35.3 |
| **Work type** |  |  |  |  |  |
|  | Sedentary | 3014 | 46.9 | 2860 | 51.3 |
|  | Standing | 2011 | 31.3 | 768 | 13.8 |
|  | Manual work | 481 | 7.5 | 1623 | 29.1 |
|  | Retired | 230 | 3.6 | 134 | 2.4 |
|  | Unemployed | 74 | 1.2 | 73 | 1.3 |
|  | Unknown | 618 | 9.6 | 116 | 2.1 |
| **Income** |  |  |  |  |  |
|  | < £20000 | 1064 | 16.6 | 552 | 9.9 |
|  | £20000 - £40000 | 2282 | 35.5 | 1853 | 33.2 |
|  | > £40000 | 2871 | 44.7 | 3057 | 54.8 |
| **Marital Status** |  |  |  |  |  |
|  | Single | 422 | 6.6 | 418 | 7.5 |
|  | Married/living as married | 3990 | 62.1 | 3605 | 64.7 |
|  | Widowed/separated/divorced | 567 | 8.8 | 309 | 5.5 |
|  | Unknown | 1449 | 22.5 | 1242 | 22.3 |
| **Smoker Status** |  |  |  |  |  |
|  | Never smoked | 3590 | 55.9 | 2871 | 51.5 |
|  | Ex smoker | 2061 | 32.1 | 1900 | 34.1 |
|  | Current smoker | 701 | 10.9 | 742 | 13.3 |
| **Site** |  |  |  |  |  |
|  | Cambridge | 2249 | 35.0 | 2096 | 37.6 |
|  | Ely | 2438 | 37.9 | 1968 | 35.3 |
|  | Wisbech | 1741 | 27.1 | 1510 | 27.1 |
| **PAEE (kJ/day/kg)** |  | 49.7 | 19.6 | 58.8 | 23.0 |
| **MVPA in bouts > 1 minute (min/day)** |  | 83.4 | 67.2 | 124.0 | 84.7 |
| **MVPA in bouts > 10 minutes (min/day)** |  | 32.9 | 39.4 | 60.3 | 58.3 |
| **Accelerometry (m/s<sup>2</sup>)** |  | 0.12 | 0.06 | 0.12 | 0.05 |

Data are mean (SD) or n (%)

Age and BMI were important correlates of PAEE, 1-minute bouted MVPA and 10-minute bouted MVPA, with levels of all three outcomes significantly lower in the oldest age group (60-64y) than the youngest age (group 29-34y), and in those classified as obese compared to normal weight participants (Table 2; p < 0.01). Figure 1 presents box plots (median and IQR) of PAEE by three age categories and three BMI groups (29-44y, 45-54y, 55-64y; normal weight, overweight, obese) and is reflective of the overall age and BMI trends with respect to PAEE. In both men and women, PAEE was highest in younger age categories and progressively lower in those 45 years and onwards. PAEE was consistently lower with advancing age than MVPA, which was not significantly different in either sex between those in their 30s and 40s, but lower in those in the 6^th^ and 7^th^ decade of life (Table 2). Similarly, those with the lowest BMI recorded the highest levels of PA, with gradually lower levels noted with higher BMI; a trend which was more pronounced in women. Sensitivity analysis conducted with allometrically scaled PAEE confirmed BMI to be a significant correlate of PAEE and MVPA, with differences between normal weight and obese participants preserved (Supplementary Table 3). Proportion of week/weekend wear was not associated with PAEE but there was a trend towards those recruited later in the study period of 2005 to 2015 having lower activity levels.

**Table 2:**
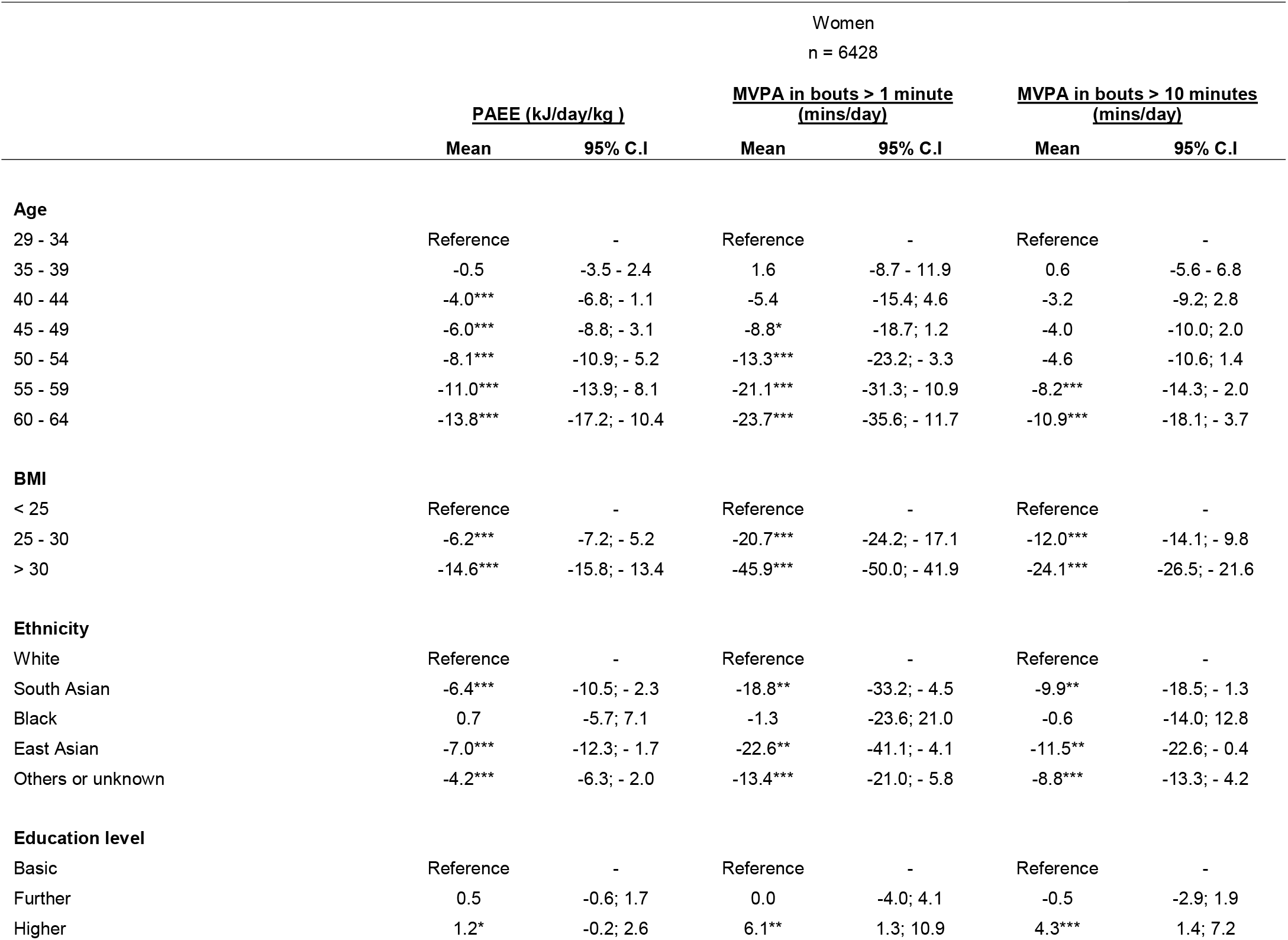
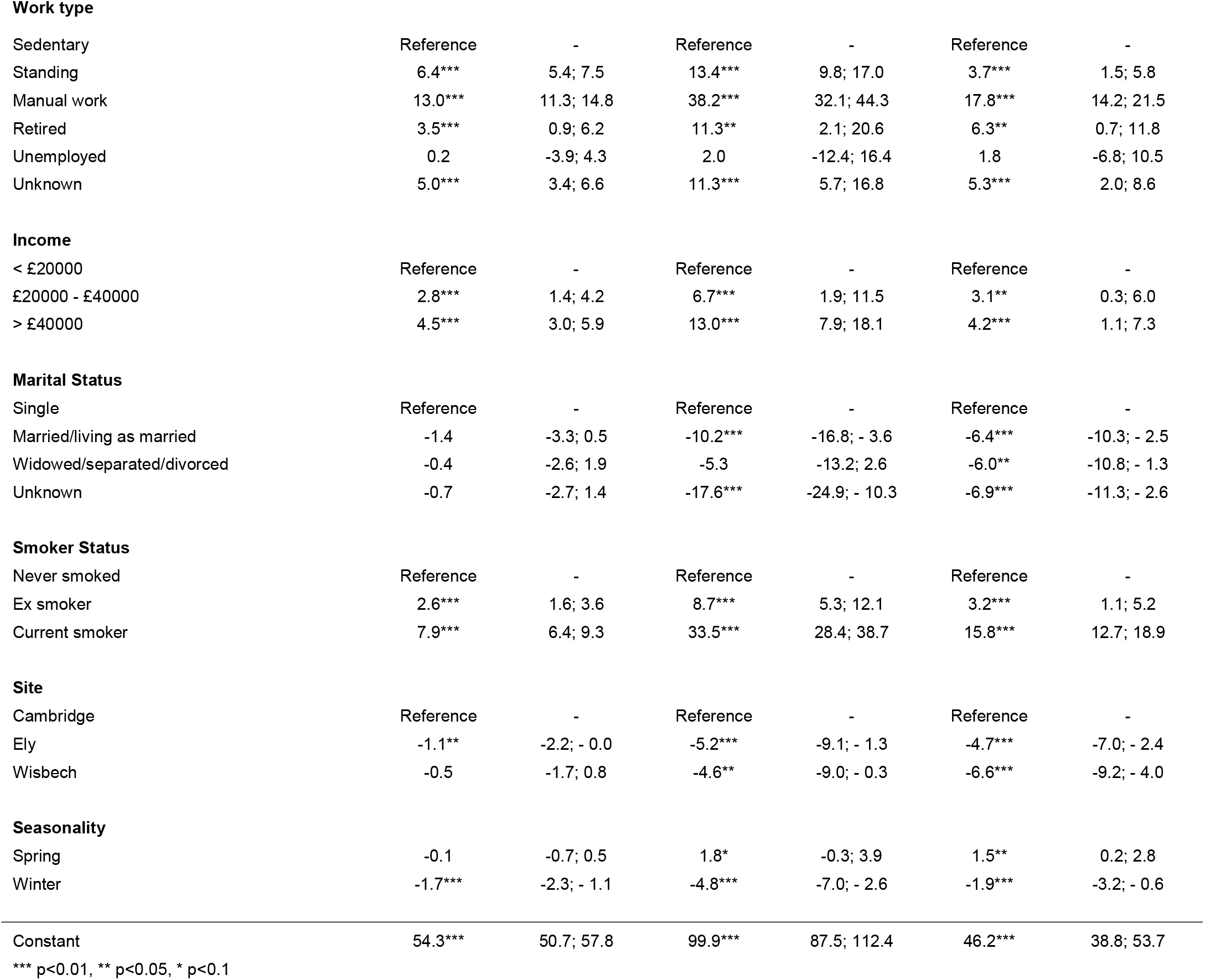
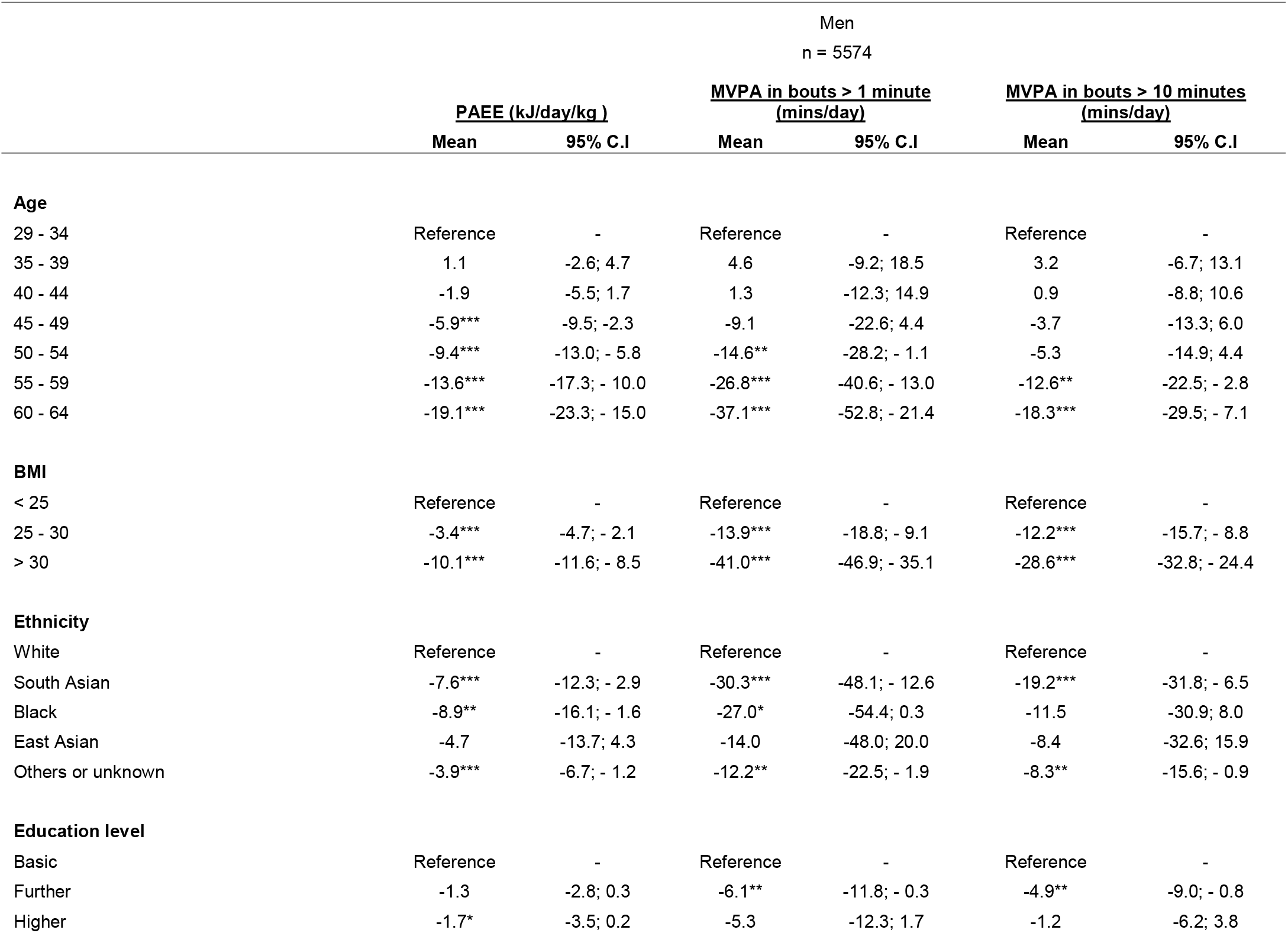
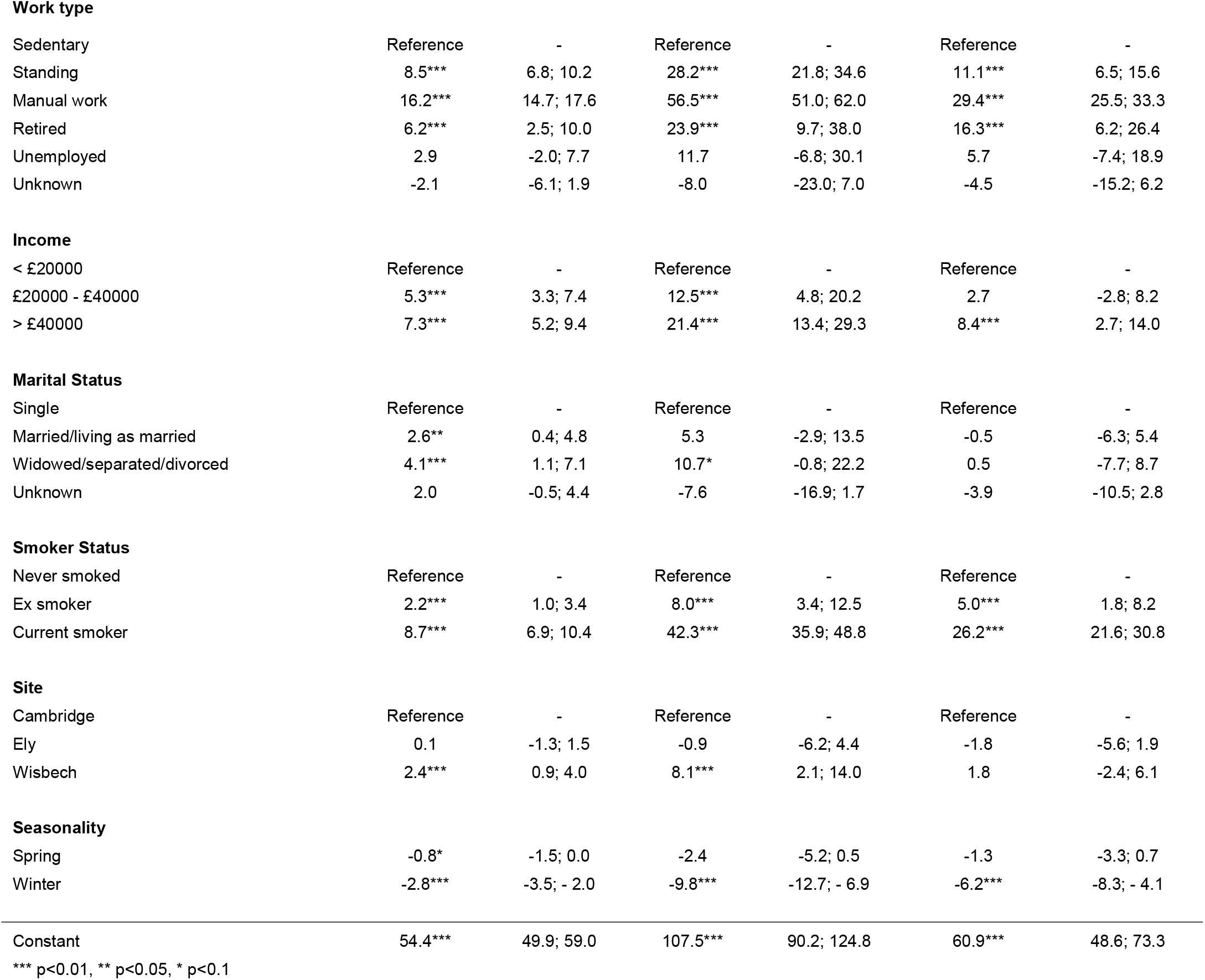
Multivariable analysis of physical activity by socio-demographic factors. The Fenland Study 2005 to 2015.

| Women<br>n = 6428 |  |  |  |  |  |  |
| --- | --- | --- | --- | --- | --- | --- |
|  | <u>PAEE (kJ/day/kg )</u> |  | <u>MVPA in bouts > 1 minute</u><br><u>(mins/day)</u> |  | <u>MVPA in bouts > 10 minutes</u><br><u>(mins/day)</u> |  |
|  | Mean | 95% C.I | Mean | 95% C.I | Mean | 95% C.I |
| **Age** |  |  |  |  |  |  |
| 29 - 34 | Reference | - | Reference | - | Reference | - |
| 35 - 39 | -0.5 | -3.5 - 2.4 | 1.6 | -8.7 - 11.9 | 0.6 | -5.6 - 6.8 |
| 40 - 44 | -4.0*** | -6.8; - 1.1 | -5.4 | -15.4; 4.6 | -3.2 | -9.2; 2.8 |
| 45 - 49 | -6.0*** | -8.8; - 3.1 | -8.8* | -18.7; 1.2 | -4.0 | -10.0; 2.0 |
| 50 - 54 | -8.1*** | -10.9; - 5.2 | -13.3*** | -23.2; - 3.3 | -4.6 | -10.6; 1.4 |
| 55 - 59 | -11.0*** | -13.9; - 8.1 | -21.1*** | -31.3; - 10.9 | -8.2*** | -14.3; - 2.0 |
| 60 - 64 | -13.8*** | -17.2; - 10.4 | -23.7*** | -35.6; - 11.7 | -10.9*** | -18.1; - 3.7 |
| **BMI** |  |  |  |  |  |  |
| < 25 | Reference | - | Reference | - | Reference | - |
| 25 - 30 | -6.2*** | -7.2; - 5.2 | -20.7*** | -24.2; - 17.1 | -12.0*** | -14.1; - 9.8 |
| > 30 | -14.6*** | -15.8; - 13.4 | -45.9*** | -50.0; - 41.9 | -24.1*** | -26.5; - 21.6 |
| **Ethnicity** |  |  |  |  |  |  |
| White | Reference | - | Reference | - | Reference | - |
| South Asian | -6.4*** | -10.5; - 2.3 | -18.8** | -33.2; - 4.5 | -9.9** | -18.5; - 1.3 |
| Black | 0.7 | -5.7; 7.1 | -1.3 | -23.6; 21.0 | -0.6 | -14.0; 12.8 |
| East Asian | -7.0*** | -12.3; - 1.7 | -22.6** | -41.1; - 4.1 | -11.5** | -22.6; - 0.4 |
| Others or unknown | -4.2*** | -6.3; - 2.0 | -13.4*** | -21.0; - 5.8 | -8.8*** | -13.3; - 4.2 |
| **Education level** |  |  |  |  |  |  |
| Basic | Reference | - | Reference | - | Reference | - |
| Further | 0.5 | -0.6; 1.7 | 0.0 | -4.0; 4.1 | -0.5 | -2.9; 1.9 |
| Higher | 1.2* | -0.2; 2.6 | 6.1** | 1.3; 10.9 | 4.3*** | 1.4; 7.2 |
| Work type |  |  |  |  |  |  |
| Sedentary | Reference | - | Reference | - | Reference | - |
| Standing | 6.4*** | 5.4; 7.5 | 13.4*** | 9.8; 17.0 | 3.7*** | 1.5; 5.8 |
| Manual work | 13.0*** | 11.3; 14.8 | 38.2*** | 32.1; 44.3 | 17.8*** | 14.2; 21.5 |
| Retired | 3.5*** | 0.9; 6.2 | 11.3** | 2.1; 20.6 | 6.3** | 0.7; 11.8 |
| Unemployed | 0.2 | -3.9; 4.3 | 2.0 | -12.4; 16.4 | 1.8 | -6.8; 10.5 |
| Unknown | 5.0*** | 3.4; 6.6 | 11.3*** | 5.7; 16.8 | 5.3*** | 2.0; 8.6 |
| Income |  |  |  |  |  |  |
| < £20000 | Reference | - | Reference | - | Reference | - |
| £20000 - £40000 | 2.8*** | 1.4; 4.2 | 6.7*** | 1.9; 11.5 | 3.1** | 0.3; 6.0 |
| > £40000 | 4.5*** | 3.0; 5.9 | 13.0*** | 7.9; 18.1 | 4.2*** | 1.1; 7.3 |
| Marital Status |  |  |  |  |  |  |
| Single | Reference | - | Reference | - | Reference | - |
| Married/living as married | -1.4 | -3.3; 0.5 | -10.2*** | -16.8; - 3.6 | -6.4*** | -10.3; - 2.5 |
| Widowed/separated/divorced | -0.4 | -2.6; 1.9 | -5.3 | -13.2; 2.6 | -6.0** | -10.8; - 1.3 |
| Unknown | -0.7 | -2.7; 1.4 | -17.6*** | -24.9; - 10.3 | -6.9*** | -11.3; - 2.6 |
| Smoker Status |  |  |  |  |  |  |
| Never smoked | Reference | - | Reference | - | Reference | - |
| Ex smoker | 2.6*** | 1.6; 3.6 | 8.7*** | 5.3; 12.1 | 3.2*** | 1.1; 5.2 |
| Current smoker | 7.9*** | 6.4; 9.3 | 33.5*** | 28.4; 38.7 | 15.8*** | 12.7; 18.9 |
| Site |  |  |  |  |  |  |
| Cambridge | Reference | - | Reference | - | Reference | - |
| Ely | -1.1** | -2.2; - 0.0 | -5.2*** | -9.1; - 1.3 | -4.7*** | -7.0; - 2.4 |
| Wisbech | -0.5 | -1.7; 0.8 | -4.6** | -9.0; - 0.3 | -6.6*** | -9.2; - 4.0 |
| Seasonality |  |  |  |  |  |  |
| Spring | -0.1 | -0.7; 0.5 | 1.8* | -0.3; 3.9 | 1.5** | 0.2; 2.8 |
| Winter | -1.7*** | -2.3; - 1.1 | -4.8*** | -7.0; - 2.6 | -1.9*** | -3.2; - 0.6 |
| Constant | 54.3*** | 50.7; 57.8 | 99.9*** | 87.5; 112.4 | 46.2*** | 38.8; 53.7 |

\*\*\* p<0.01, \*\* p<0.05, \* p<0.1

|  | Men<br>n = 5574 |  |  |  |  |  |
| --- | --- | --- | --- | --- | --- | --- |
|  | <u>PAEE (kJ/day/kg )</u> |  | <u>MVPA in bouts > 1 minute</u><br><u>(mins/day)</u> |  | <u>MVPA in bouts > 10 minutes</u><br><u>(mins/day)</u> |  |
|  | Mean | 95% C.I | Mean | 95% C.I | Mean | 95% C.I |
| **Age** |  |  |  |  |  |  |
| 29 - 34 | Reference | - | Reference | - | Reference | - |
| 35 - 39 | 1.1 | -2.6; 4.7 | 4.6 | -9.2; 18.5 | 3.2 | -6.7; 13.1 |
| 40 - 44 | -1.9 | -5.5; 1.7 | 1.3 | -12.3; 14.9 | 0.9 | -8.8; 10.6 |
| 45 - 49 | -5.9*** | -9.5; -2.3 | -9.1 | -22.6; 4.4 | -3.7 | -13.3; 6.0 |
| 50 - 54 | -9.4*** | -13.0; - 5.8 | -14.6** | -28.2; - 1.1 | -5.3 | -14.9; 4.4 |
| 55 - 59 | -13.6*** | -17.3; - 10.0 | -26.8*** | -40.6; - 13.0 | -12.6** | -22.5; - 2.8 |
| 60 - 64 | -19.1*** | -23.3; - 15.0 | -37.1*** | -52.8; - 21.4 | -18.3*** | -29.5; - 7.1 |
| **BMI** |  |  |  |  |  |  |
| < 25 | Reference | - | Reference | - | Reference | - |
| 25 - 30 | -3.4*** | -4.7; - 2.1 | -13.9*** | -18.8; - 9.1 | -12.2*** | -15.7; - 8.8 |
| > 30 | -10.1*** | -11.6; - 8.5 | -41.0*** | -46.9; - 35.1 | -28.6*** | -32.8; - 24.4 |
| **Ethnicity** |  |  |  |  |  |  |
| White | Reference | - | Reference | - | Reference | - |
| South Asian | -7.6*** | -12.3; - 2.9 | -30.3*** | -48.1; - 12.6 | -19.2*** | -31.8; - 6.5 |
| Black | -8.9** | -16.1; - 1.6 | -27.0* | -54.4; 0.3 | -11.5 | -30.9; 8.0 |
| East Asian | -4.7 | -13.7; 4.3 | -14.0 | -48.0; 20.0 | -8.4 | -32.6; 15.9 |
| Others or unknown | -3.9*** | -6.7; - 1.2 | -12.2** | -22.5; - 1.9 | -8.3** | -15.6; - 0.9 |
| **Education level** |  |  |  |  |  |  |
| Basic | Reference | - | Reference | - | Reference | - |
| Further | -1.3 | -2.8; 0.3 | -6.1** | -11.8; - 0.3 | -4.9** | -9.0; - 0.8 |
| Higher | -1.7* | -3.5; 0.2 | -5.3 | -12.3; 1.7 | -1.2 | -6.2; 3.8 |
| **Work type** |  |  |  |  |  |  |
| Sedentary | Reference | - | Reference | - | Reference | - |
| Standing | 8.5*** | 6.8; 10.2 | 28.2*** | 21.8; 34.6 | 11.1*** | 6.5; 15.6 |
| Manual work | 16.2*** | 14.7; 17.6 | 56.5*** | 51.0; 62.0 | 29.4*** | 25.5; 33.3 |
| Retired | 6.2*** | 2.5; 10.0 | 23.9*** | 9.7; 38.0 | 16.3*** | 6.2; 26.4 |
| Unemployed | 2.9 | -2.0; 7.7 | 11.7 | -6.8; 30.1 | 5.7 | -7.4; 18.9 |
| Unknown | -2.1 | -6.1; 1.9 | -8.0 | -23.0; 7.0 | -4.5 | -15.2; 6.2 |
| **Income** |  |  |  |  |  |  |
| < £20000 | Reference | - | Reference | - | Reference | - |
| £20000 - £40000 | 5.3*** | 3.3; 7.4 | 12.5*** | 4.8; 20.2 | 2.7 | -2.8; 8.2 |
| > £40000 | 7.3*** | 5.2; 9.4 | 21.4*** | 13.4; 29.3 | 8.4*** | 2.7; 14.0 |
| **Marital Status** |  |  |  |  |  |  |
| Single | Reference | - | Reference | - | Reference | - |
| Married/living as married | 2.6** | 0.4; 4.8 | 5.3 | -2.9; 13.5 | -0.5 | -6.3; 5.4 |
| Widowed/separated/divorced | 4.1*** | 1.1; 7.1 | 10.7* | -0.8; 22.2 | 0.5 | -7.7; 8.7 |
| Unknown | 2.0 | -0.5; 4.4 | -7.6 | -16.9; 1.7 | -3.9 | -10.5; 2.8 |
| **Smoker Status** |  |  |  |  |  |  |
| Never smoked | Reference | - | Reference | - | Reference | - |
| Ex smoker | 2.2*** | 1.0; 3.4 | 8.0*** | 3.4; 12.5 | 5.0*** | 1.8; 8.2 |
| Current smoker | 8.7*** | 6.9; 10.4 | 42.3*** | 35.9; 48.8 | 26.2*** | 21.6; 30.8 |
| **Site** |  |  |  |  |  |  |
| Cambridge | Reference | - | Reference | - | Reference | - |
| Ely | 0.1 | -1.3; 1.5 | -0.9 | -6.2; 4.4 | -1.8 | -5.6; 1.9 |
| Wisbech | 2.4*** | 0.9; 4.0 | 8.1*** | 2.1; 14.0 | 1.8 | -2.4; 6.1 |
| **Seasonality** |  |  |  |  |  |  |
| Spring | -0.8* | -1.5; 0.0 | -2.4 | -5.2; 0.5 | -1.3 | -3.3; 0.7 |
| Winter | -2.8*** | -3.5; - 2.0 | -9.8*** | -12.7; - 6.9 | -6.2*** | -8.3; - 4.1 |
| Constant | 54.4*** | 49.9; 59.0 | 107.5*** | 90.2; 124.8 | 60.9*** | 48.6; 73.3 |

\*\*\* p<0.01, \*\* p<0.05, \* p<0.1

**Figure 1:**
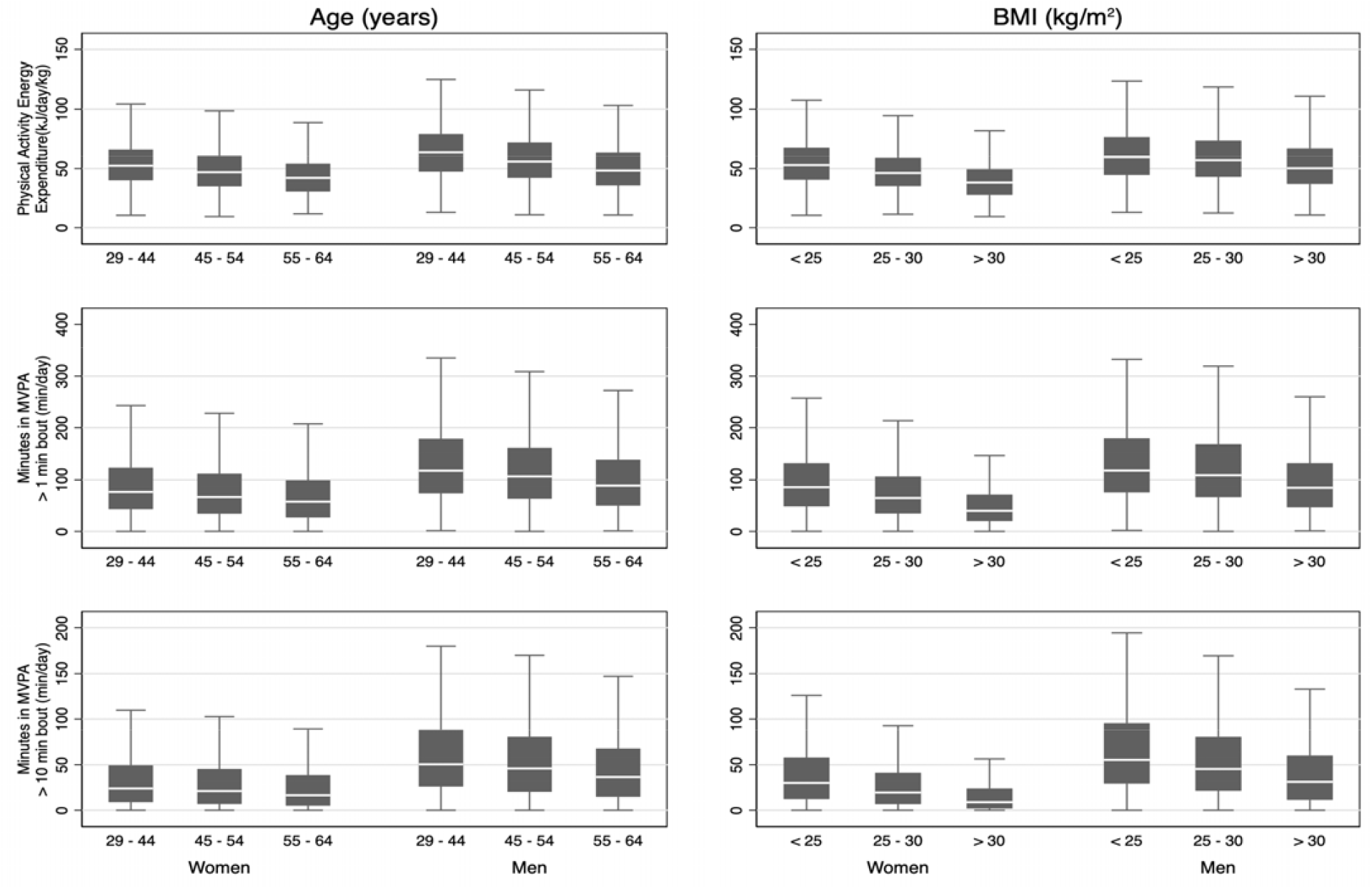
Sex stratified physical activity energy expenditure (PAEE) in kJ/day/kg by age and BMI in the Fenland cohort. Men accumulated higher levels of PAEE than women, with younger participants accumulating more PA regardless of sex. PAEE is inversely related to BMI. Box plots represent medians and interquartile ranges.

Across the sample, LPA was the primary contributor of energy to total PAEE (59% women, 51% men), although men accumulated higher levels of MPA and VPA than women (Figure 2). However, the majority of time was spent asleep or sedentary (70% women, 69% men), demonstrating the paradox that participants spent a relatively small fraction of the day accumulating most of their PAEE. Women spent almost all their time below 9 METs and men below 11 METS (Supplementary Figure 1). Time use demonstrated a similar association with age as for PAEE; time in LPA was lower in older participants, with a reciprocal higher time spent asleep or sedentary. This was accompanied by a commensurate lower PAEE derived from LPA in older participants.

**Figure 2:**
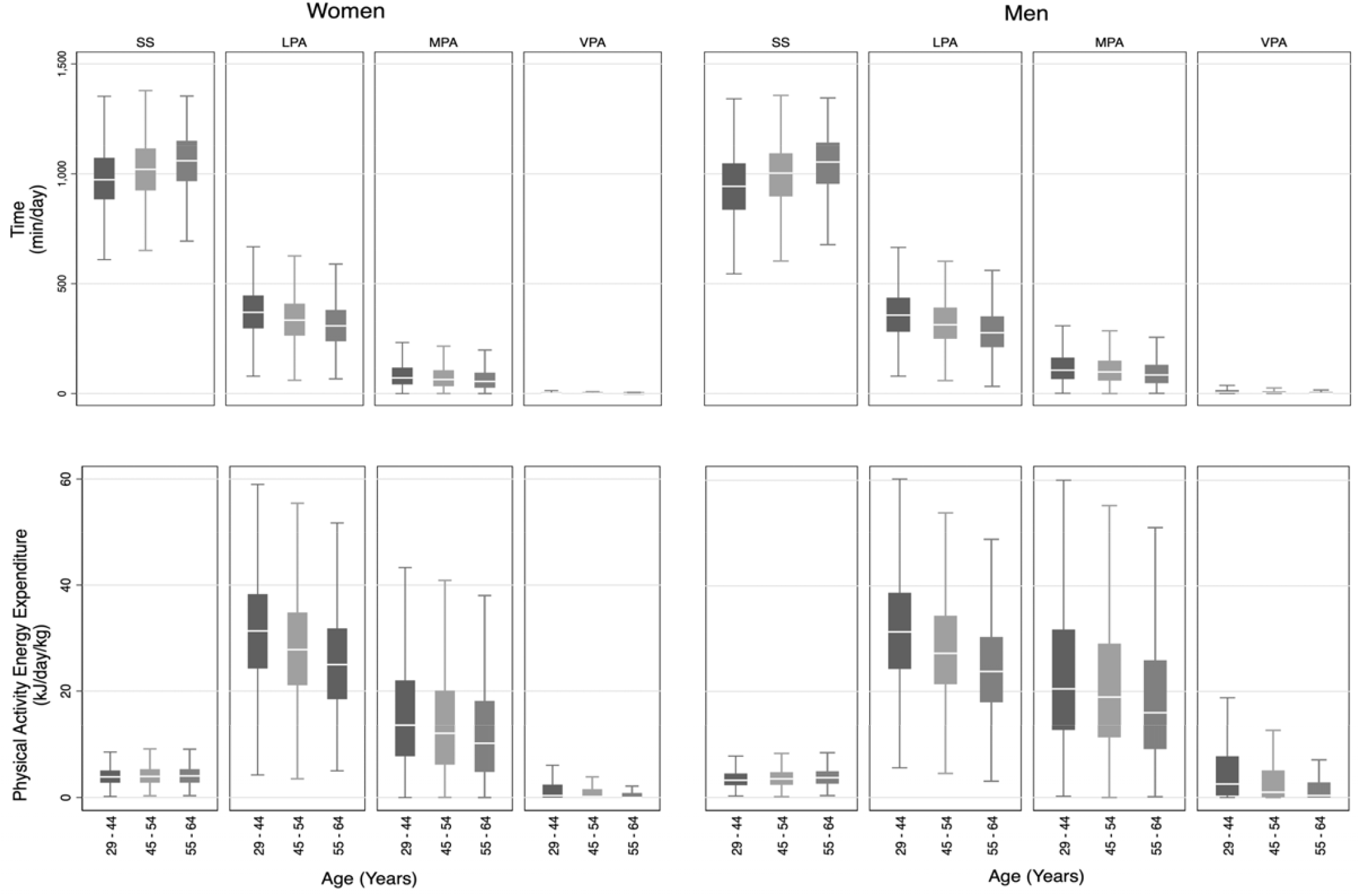
Sex stratified intensities of physical activity by time (minutes per day) and energy expenditure (kJ/day/kg) in the Fenland cohort. SS = sedentary or sleep (<1.5 METs); LPA = light physical activity (1.5-3METs); MPA = moderate physical activity (3-6METs); VPA = vigorous physical activity (>6METs) Box plots represent medians and interquartile ranges.

Multivariable analysis (Table 2) showed further correlates of PA, including ethnicity, work-type, income, and smoker status. Ethnic minorities tended to be less active than White participants, although the sample sizes of some groups are too small to assess this with certainty. Participants with more physically demanding jobs also had higher activity levels; however retired individuals were more active than those with sedentary occupations. When adjusted for all other socioeconomic variables, this equated to women in manual occupations accumulating 13 kJ•day^-1^•kg^-1^ more PAEE than those in sedentary jobs, and 38 and 18 more minutes of MVPA in bouts of >1 minute and >10 minutes respectively. In men, corresponding differences were 16 kJ•day^-1^•kg^-1^, 56 and 29 min/day.

Education was not significantly associated with PA after adjustment (Table 2). However, there was some indication of higher activity levels in those with higher socio-economic status, more so in men for income level. This picture was mixed as demonstrated by the higher activity in men and trend towards lower activity in women from the Wisbech site, compared to the more affluent Cambridge site. A sex difference was also observed for civil status, in that marriage (current or former) was positively associated with activity in men but tended to be inversely associated in women, compared to single status. Current and former smokers tended to have higher PAEE and MVPA than non-smokers for both sexes; however, smokers moved less when only considering the accelerometry component (Supplementary Table 2). Participants who were measured in the summer were more active than those measured in the winter; this difference was twice as big in men as in women, i.e. 20-min and 10-min difference in 1-minute bouted MVPA between mid-winter and mid-summer.

Overall, compliance with the current UK PA guidelines (as defined by 150 minutes of MVPA per week in bouts > 10minutes) was higher in men than in women; 49% of women and 74% of men accumulated at least 21.4 min of MVPA/day (equivalent to meeting the guidelines of 150 min MVPA per week) in bouts lasting 10-min or longer when MVPA was defined as > 3 METs. When redefined at MVPA > 3 METs of at least 1-minute duration, however, compliance was 86% and 96%, respectively. Conversely, redefining MVPA with the higher 4 METs cut-off naturally resulted in lower absolute levels of MVPA and guideline compliance, as highlighted in Figure 3. Using a 4 MET threshold and 10-minute bout definition, only 15% of women and 31% of men accumulated the recommended levels of MVPA. Participants accumulated less than half the amount of 10-minute bouted MVPA than 1-minute bouted MVPA, irrespective of sex or MVPA cut-offs. Using the stricter definition of MVPA, sensitivity analyses of the association with sociodemographic factors (Supplementary Table 1) showed largely similar patterns of association to the primary analysis.

**Figure 3:**
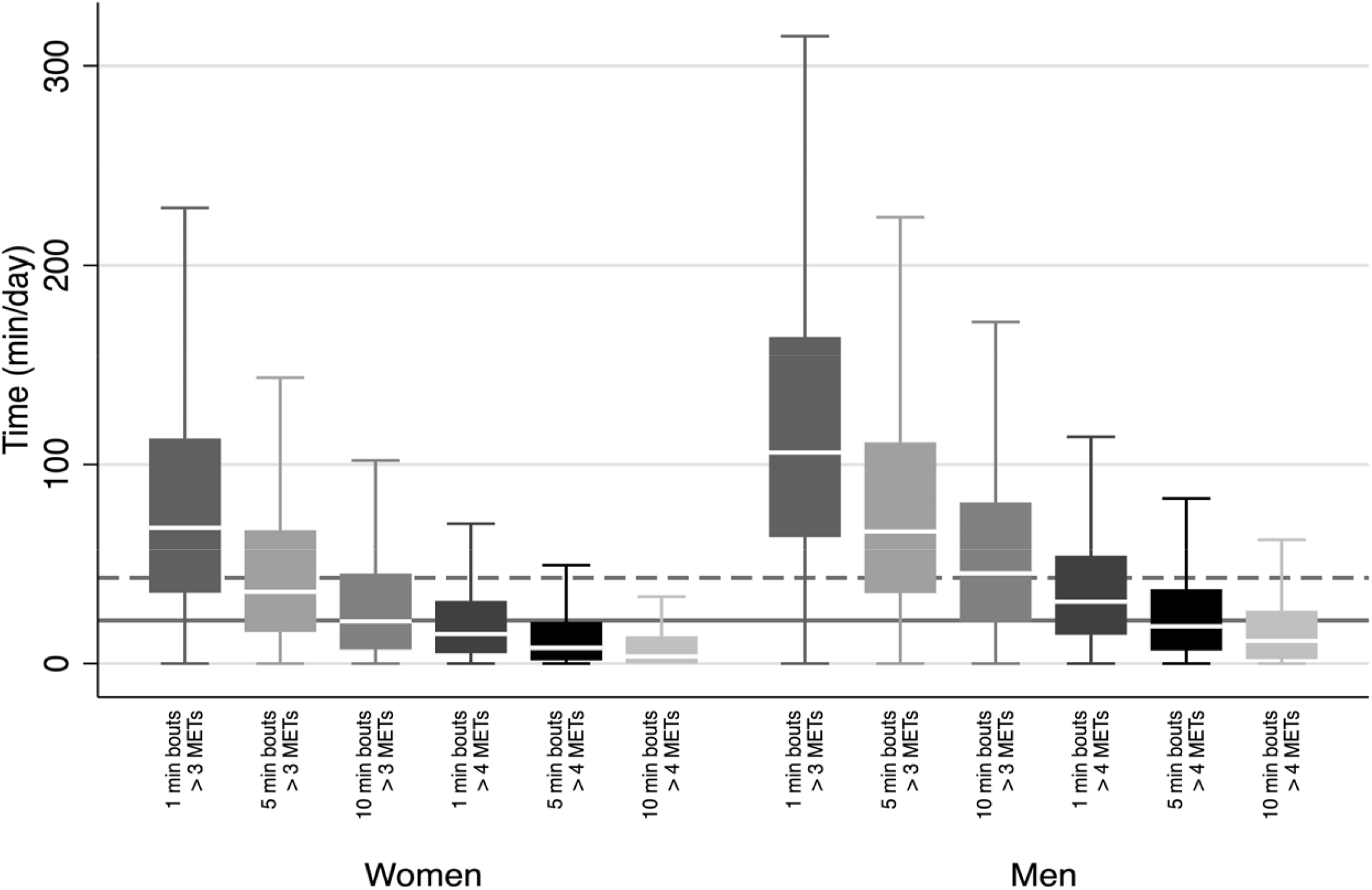
Sex stratified moderate to vigorous physical activity (MVPA) of differing bout lengths, according to 2 intensity thresholds in the Fenland cohort. The solid grey line indicates compliance with UK physical activity guidelines, the dashed line represents double the UK guidelines. Box plots represent medians and interquartile ranges.

## Discussion

In this study we present the descriptive epidemiology of physical activity in a population-sample of UK adults. To our knowledge, this is the first study that describes socio-demographic patterns of both PAEE and its underlying intensity distribution in a large cohort of younger to middle-aged adults.

Our results indicate that men accumulate higher levels of PAEE than women across all ages; a finding consistent with other descriptive studies [7, 22–24]. Men also accumulate higher levels of MVPA and expend more of their energy at higher intensities than women. Despite the energy expenditure differences between men and women as measured by combined sensing, there was no sex difference in movement as measured by accelerometry, as has also been observed in other cohorts [25].

Among both men and women PA was lower in older individuals. Moreover, the levels and age-associated differences in PAEE demonstrated in Figure 1 are similar to that recently reported in UK nationally representative samples [7], and the age association remained significant in multivariable analysis (Table 2). Although PAEE and MVPA were highest in young adults, the pattern of association with age was quite different. Whereas PAEE was significantly different between all but the youngest two groups in women and youngest three groups in men, MVPA in >1-min bouts was only significantly different between the reference groups and those above 50y (p < 0.05). Furthermore, MVPA in 10-min bouts was only significantly different between the youngest group and the oldest two age groups in both sexes. This finding suggests that intensity patterns of PA may be preserved differently as people age, with light LPA replaced by sedentary behaviour proportionally more rapidly than decreases in MVPA.

Normal-weight individuals accumulated higher levels of all activity outcomes than obese individuals. The difference in PAEE between normal weight and obese women was similar to the difference between sedentary and manual workers and between participants in the oldest and youngest age categories. The same comparisons hold true for men. This finding was consistent in a sensitivity analysis for allometrically scaled PAEE (Supplementary Table 3), suggesting that obesity is negatively associated with levels of PA in both sexes, irrespective of the effect of body size on absolute energy expenditure.

In terms of socio-demographic correlates of PAEE and MVPA, our unadjusted results demonstrated that educational level, income, work type, and smoking status were associated with PAEE and MVPA in women and men. Yet, when mutually adjusted in the multivariable analysis, some of these differences were attenuated. For example, univariable analysis showed a 16 kJ•day^-1^•kg^-1^ difference between men with sedentary jobs and men with manual jobs. Likewise, univariable analysis modelled a 6 kJ•day^-1^•kg^-1^ difference between men with a basic education compared to those with higher education. When controlled for all sociodemographic covariates, the association between PAEE and education in men decreased in magnitude and significance, whereas the association with different work types remained significant, and of an order of magnitude almost identical to univariable analysis. A similar trend was evident in women. This is due to the strong correlations between certain sociodemographic variables such as lower levels of education and manual work. Conversely, higher levels of education tended to be associated with less PA (in men) but must be balanced against the higher levels of PAEE and MVPA seen with increasing income.

Similarly, the higher attributed levels of PAEE in smokers should be considered in the context of significantly lower accelerometry-measured PA in current smokers of both sexes in multivariable analysis (Supplementary Table 3). Smoking is known to increase energy expenditure, independent of movement and resting energy expenditure [26]. Moreover, this combination of results may be a partial by-product of the acute increase in heart rate of 15 beats/min that has been observed in cigarette smokers [27], which may have biased the estimates of energy expenditure.

The levels of PAEE in the Fenland cohort are comparable to those reported in the nationally representative sample of the UK population studied with the gold-standard method of doubly labelled water in NDNS [7]. That study reported mean (SD) PAEE of 52 (20) and 47 (17) kJ•day^-1^•kg^-1^ in women aged 16–49 and 50–64 respectively. By contrast, men accumulated 63 (23) and 54 (20) kJ•day^-1^•kg^-1^ in the same age groups. The Cambridge-based ROOTS study reported higher PAEE levels of 83.5 and 65.9 kJ•day^-1^•kg^-1^ in 825, 16-year old boys and girls respectively [8]. By contrast, Golubic et al reported comparatively lower PAEE levels in the nationally representative 1946 birth cohort of British women and men assessed at age 60-64 years (median (IQR) PAEE: 33.5 and 35.5 kJ•day^-1^•kg^-1^ respectively) [9],suggesting possible regional differences within the UK.

Comparing the Fenland cohort to populations living in locations of differing global developmental indices, we note that various rural cohorts in Africa are more active. However, while men living in urban environments in developing countries have comparable levels of PA to their Cambridgeshire counterparts, women are notably less active. For example, Kenyan men of Kamba, Maasai and Luo ethnic origins had PAEE levels of 81, 78 and 74 kJ•day^-1^•kg^-1^ (age-adjusted to 40 years), respectively [23]. In contrast, Cameroonian and Barbadian urban men recorded comparable levels of PAEE to Fenland men (Cameroon: 52 kJ•day^-1^•kg^-1^, 37-years-old; Barbadian: 47 kJ•day^-1^•kg^-1^, 25-54 years-old) whereas Fenland women were more active than the women in those populations (Cameroon: 38 kJ•day^-1^•kg^-1^, 38-years-old; Barbadian: 36 kJ•day^-1^•kg^-1^, 25-54 years-old) [22, 24]. This suggests different sex-specific determinants of PAEE in the developed as opposed to the developing world, and rural compared to urbanised environments.

Our study also demonstrates how the definition of MVPA influences the reported prevalence of compliance with physical activity guidelines. At a time when PA guidelines are undergoing review [28], the difference in the quantity of MVPA accumulated in bouts of at least 1-min duration vs at least 10-min duration is critical. Through the simple abolition of the 10-min bouted criterion, apparent compliance falls from 86% to 49% in women and 96% to 74% in men (Figure 3). This difference reflects characterisation according to the new US guidelines [4], as opposed to the 2010 UK guidelines [5]. Furthermore, the new US guidelines emphasise the role of all physical activity in health. Our study suggests that LPA plays a crucial role in the accumulation of overall PAEE, accounting for roughly two thirds of PAEE in women and over half of PAEE in men (Figure 2). Indeed, without reasonable levels of LPA, it is hard to accumulate high levels of PAEE. Therefore, LPA’s role in health should not be underestimated.

Strengths of this study include individually calibrated combined heart rate and movement-based assessment of PAEE and underlying intensity in a large population-based sample that compares well in terms of PAEE with a nationally representative sample measured by gold-standard measures (i.e. NDNS)[7]. The large sample allowed for detailed description of physical activity by several sociodemographic variables. A further strength is our ability to quantify potential selection bias by comparison of the recruited cohort to the sampling frame population in Cambridgeshire. Whilst this does suggest that the participants are slightly healthier (lower BMI, smoke less, and drink fewer units of alcohol per week) and live in less deprived areas, the differences are small and their likely impact on quantified activity levels not always in the same direction, hence some of the bias will cancel out. Limitations of the study include its lack of representativeness compared to the rest of the UK, in particular the predominance of White participants.

## Conclusions

In this study we have described the objectively measured physical activity levels of young to middle-aged adults in the East of England, which are similar to those reported in the nationally representative NDNS study [7]. As with other British studies age, sex and BMI were important correlates of physical activity. We have additionally shown work type, income and smoker status to be associated with MVPA and energy expenditure. Overall, 86% of women and 96% of men met the most liberal criteria for the PA guidelines compared to 15% and 31% respectively for the strictest. Uniquely, we have identified LPA as the main driver of PAEE; a component of PA that is currently not quantified as a target in UK guidelines.

## Data Availability

The data used for this study is available upon request via the link below.

http://www.mrc-epid.cam.ac.uk/research/data-sharing/

## List of abbreviations

BMI: Body mass index
LPA: Light physical activity
MET(s): Metabolic equivalent of task
MPA: Moderate physical activity
MVPA: Moderate-to-vigorous physical activity
NDNS: National Diet and Nutrition Survey
PA: Physical activity
PAEE: Physical activity energy expenditure
SS: Sedentary and sleep
VPA: Vigorous physical activity

## Declarations

### Ethics approval and consent to participate

All participants provided written informed consent and the study was approved by the local ethics committee (NRES Committee – East of England Cambridge Central) and performed in accordance with the Declaration of Helsinki.

## Consent for publication

Not applicable

## Availability of data and material

The datasets generated and analysed during the current study are available at request via the MRC Epidemiology website (http://www.mrc-epid.cam.ac.uk/research/data-sharing/)

## Authors’ contributions

The authors contributed to the present manuscript as follows: Idea for analysis (TL, SB); acquisition, analysis of raw physical activity data (KW, SH, SB); acquisition, analysis of General Practice data (NK, TL), epidemiological data analysis (TL); drafting of the manuscript (TL); revising work critically for important intellectual content (all authors); approval of the final version before submission (all authors). Chief Investigator (NJW) and Principal Investigators (NF, SG, SB) of the Fenland Study.

## Acknowledgements

We are grateful to the Fenland Study participants for their willingness and time to take part. We thank all members of the following teams responsible for practical aspects of the study; Study Coordination, Field Epidemiology, Anthropometry Team, Physical Activity Technical Team, IT, Data Management, and Statistics.

## Table and figure captions

**Supplementary Table 1:** Comparison of Fenland sampling frame and study subpopulation. The Fenland Study 2005 to 2015.

**Supplementary Table 2:** Univariable analysis of physical activity by socio-demographic factors in women. The Fenland Study 2005 to 2015.

**Supplementary Table 3:** Multivariable analysis of socio-demographic factors in men using accelerometry counts, allometric scaling of body weight for PAEE, and a higher intensity criterion for MVPA. The Fenland Study 2005 to 2015.

**Supplementary Figure 1:** Sex stratified time (minutes per day) across the full spectrum of physical activity intensities in the Fenland cohort. Men accumulated higher levels of very vigorous physical activity than women, although both sexes accumulated relatively little activity over 5 METs. Box plots represent medians and interquartile ranges.

## References

1. Lee I-M, Shiroma EJ, Lobelo F, Puska P, Blair SN, Katzmarzyk PT, et al. Effect of physical inactivity on major non-communicable diseases worldwide: an analysis of burden of disease and life expectancy. Lancet. 2012;380:219–29. doi:10.1016/S0140-6736(12)61031-9.

2. Smith AD, Crippa A, Woodcock J, Brage S. Physical activity and incident type 2 diabetes mellitus: a systematic review and dose–response meta-analysis of prospective cohort studies. Diabetologia. 2016;59:2527–45. doi:10.1007/s00125-016-4079-0.

3. Hansen A-LS, Carstensen B, Helge JW, Johansen NB, Gram B, Christiansen JS, et al. Combined Heart Rate– and Accelerometer-Assessed Physical Activity Energy Expenditure and Associations With Glucose Homeostasis Markers in a Population at High Risk of Developing Diabetes. Diabetes Care. 2013;36:3062–9. doi:10.2337/DC12-2671.

4. Piercy KL, Troiano RP, Ballard RM, Carlson SA, Fulton JE, Galuska DA, et al. The Physical Activity Guidelines for Americans. JAMA. 2018;320:2020. doi:10.1001/jama.2018.14854.

5. Bull FC, The Expert Working Groups. Physical Activity Guidelines in the UK: Review and Recommendations. Sch Sport Exerc Heal Sci Loughbrgh Univ. 2010. doi:10.1071/EA03155.

6. Van Remoortel H, Camillo CA, Langer D, Hornikx M, Demeyer H, Burtin C, et al. Moderate intense physical activity depends on selected Metabolic Equivalent of Task (MET) cut-off and type of data analysis. PLoS One. 2013;8:e84365. doi:10.1371/journal.pone.0084365.

7. Brage S, Lindsay T, Venables M, Wijndaele K, Westgate K, Collins D, et al. Descriptive epidemiology of energy expenditure in the UK: Findings from the National Diet and Nutrition Survey 2008 to 2015. bioRxiv. 2019;:542613. doi:10.1101/542613.

8. Collings PJ, Wijndaele K, Corder K, Westgate K, Ridgway CL, Dunn V, et al. Levels and patterns of objectively-measured physical activity volume and intensity distribution in UK adolescents: the ROOTS study. Int J Behav Nutr Phys Act. 2014;11:23. doi:10.1186/1479-5868-11-23.

9. Golubic R, Martin KR, Ekelund U, Hardy R, Kuh D, Wareham N, et al. Levels of physical activity among a nationally representative sample of people in early old age: results of objective and self-reported assessments. Int J Behav Nutr Phys Act. 2014;11:58. doi:10.1186/1479-5868-11-58.

10. Brage S, Brage N, Franks PW, Ekelund U, Wareham NJ. Reliability and validity of the combined heart rate and movement sensor Actiheart. Eur J Clin Nutr. 2005;59:561. https://doi.org/10.1038/sj.ejcn.1602118.

11. Brage S, Ekelund U, Brage N, Hennings MA, Froberg K, Franks PW, et al. Hierarchy of individual calibration levels for heart rate and accelerometry to measure physical activity. J Appl Physiol. 2007;103:682–92. doi:10.1152/japplphysiol.00092.2006.

12. Tanaka H, Monahan KD, Seals DR. Age-Predicted Maximal Heart Rate Revisited. 2001. doi:10.1016/S0735-1097(00)01054-8.

13. Stegle O, Fallert S V, on … DJC. Gaussian process robust regression for noisy heart rate data. Gaussian Process robust Regres noisy Hear rate data. 2008. http://scholar.google.com/scholar?q=Gaussian process robust regression for noisy heart rate data&btnG=&hl=en&num=20&as_sdt=0%2C22 VN-readcube.com.

14. Brage S, Brage N, Franks PW, Ekelund U, Wong M-Y, Andersen LB, et al. Branched equation modeling of simultaneous accelerometry and heart rate monitoring improves estimate of directly measured physical activity energy expenditure. J Appl Physiol. 2004;96:343–51. doi:10.1152/japplphysiol.00703.2003.

15. Strath SJ, Brage S, Ekelund U. Integration of physiological and accelerometer data to improve physical activity assessment. Med Sci Sports Exerc. 2005;37 11 Suppl:S563–71. http://www.ncbi.nlm.nih.gov/pubmed/16294119. Accessed 11 Mar 2019.

16. Thompson D, Batterham AM, Bock S, Robson C, Stokes K. Assessment of Low-to-Moderate Intensity Physical Activity Thermogenesis in Young Adults Using Synchronized Heart Rate and Accelerometry with Branched-Equation Modeling. J Nutr. 2006;136:1037–42. doi:10.1093/jn/136.4.1037.

17. Stegle O, Fallert S V., MacKay DJC, Brage S. Gaussian process robust regression for noisy heart rate data. IEEE Trans Biomed Eng. 2008;55:2143–51. doi:10.1109/TBME.2008.923118.

18. Brage S, Westgate K, Wijndaele K, Godinho J, Griffin S, Wareham N. Evaluation of a method for minimising diurnal information bias in objective sensor data. In: ICAMPAM (Amherst). 2013.

19. Brage S, Westgate K, Franks PW, Stegle O, Wright A, Ekelund U, et al. Estimation of free-living energy expenditure by heart rate and movement sensing: A doubly-labelled water study. PLoS One. 2015;10:1–19. doi:10.1371/journal.pone.0137206.

20. White CR, Seymour RS, Thompson MB. Allometric scaling of mammalian metabolism. J Exp Biol. 2005;208 Pt 9:1611–9. doi:10.1242/jeb.01501.

21. Sarrus M, Rameaux J. Rapport sur un memoire adresse a l’academie royale de medicine. Bull l’Academie R Med Paris. 1883;3:1094–100. http://ci.nii.ac.jp/naid/10025722524/en/. Accessed 18 Dec 2018.

22. Assah F, Mbanya JC, Ekelund U, Wareham N, Brage S. Patterns and correlates of objectively measured free-living physical activity in adults in rural and urban Cameroon. 2015;69:700–7. http://www.ncbi.nlm.nih.gov/pubmed/25841243. Accessed 9 Jan 2019.

23. Christensen DL, Faurholt-Jepsen D, Boit MK, Mwaniki DL, Kilonzo B, Tetens I, et al. Cardiorespiratory fitness and physical activity in Luo, Kamba, and Maasai of rural Kenya. Am J Hum Biol. 2012;24:723–9. doi:10.1002/ajhb.22303.

24. Howitt C, Brage S, Hambleton IR, Westgate K, Samuels TA, Rose AM, et al. A cross-sectional study of physical activity and sedentary behaviours in a Caribbean population: combining objective and questionnaire data to guide future interventions. 2016. doi:10.1186/s12889-016-3689-2.

25. Doherty A, Jackson D, Hammerla N, Plötz T, Olivier P, Granat MH, et al. Large scale population assessment of physical activity using wrist worn accelerometers: The UK biobank study. PLoS One. 2017;12:1–14.

26. Hofstetter A, Schutz Y, Jéquier E, Wahren J. Increased 24-Hour Energy Expenditure in Cigarette Smokers. N Engl J Med. 1986;314:79–82. doi:10.1056/NEJM198601093140204.

27. Houlihan ME, Pritchard WS, Robinson JH. A double blind study of the effects of smoking on heart rate: is there tachyphylaxis? Psychopharmacology (Berl). 1999;144:38–44. http://www.ncbi.nlm.nih.gov/pubmed/10379622. Accessed 3 May 2019.

28. Murphy MH, Broom DR, Gill JMR, Gray CM, Jones A, Steele J, et al. UK physical activity guidelines: Draft review and recommendations for adults (aged 19-64 years). 2018. http://www.bristol.ac.uk/media-library/sites/sps/documents/cmo/adults-technical-report.pdf. Accessed 2 Jul 2019.

